# Advanced Nursing Practice in the context of Primary Health Care in Brazil: documentary research

**DOI:** 10.1101/2025.04.30.25326567

**Authors:** Emerson Willian Santos de Almeida, Italo Rodolfo Silva, Simone de Godoy, Beatriz Rosana Gonçalves de Oliveira Toso, Elizimara Ferreira Siqueira, Ellen Marcia Peres, Jeanne-Marie Rodrigues Stacciarini, Isabel Amélia Costa Mendes

**Affiliations:** University of São Paulo. Ribeirão Preto College of Nursing, Brazil; Federal University of Rio de Janeiro. Macaé, Rio de Janeiro, Brazil; State University of Western Paraná. Cascavel, Paraná, Brazil; Primary institution. Santa Catarina, State, Brazil; State University of Rio de Janeiro. Rio de Janeiro, Rio de Janeiro, Brazil; University of Michigan. Ann Arbor, Michigan, United States

**Keywords:** Advanced Nursing Practice, Advanced Practice Nurse, Primary Health Care, Unified Health System, Nursing, Health Policy

## Abstract

**Objective:** To categorize Advanced Nursing Practice activities in Brazil, registered in the Health Production reporting platform of the Health Information System for Primary Care, by nurses from Primary Health Care.

**Methods:** a documentary, exploratory, nationwide study conducted in 2024, based on public data available on websites linked to the Ministry of Health in Brazil, reference year in 2023.

**Results:** the analysis matrix represents a comparative conceptual structure that demonstrates the evidence of advanced practices that have been assumed and recorded by nurses in the SUS data system, with open access.

**Final considerations:** The study shows that, despite the autonomy of nurses in practices such as medication prescription, test request, referral, IUD insertion, suturing and other interventions, the Brazilian policy still presents discrepancies in relation to the ICN and WHO guidelines.

## INTRODUCTION

Based on the principles of universality, comprehensiveness and equity, the Unified Health System (SUS), the health system in force in Brazil, guarantees care to the population and has been implemented for 35 years, presenting strengths and challenges (1). In this context, integrated by political, economic and institutional interactions, the understanding is corroborated that it is up to health systems to manage and provide resources to their subsystems and processes, capable of meeting the purposes in favor of the health of the population, materializing in structures, norms and services aimed at achieving results aligned with the conception of health in the context of a nation. Furthermore, its scope involves organizations, professionals, services, inputs, technologies and knowledge, with the objective of to promote health, prevent diseases and offer care and rehabilitation(2).

The relevance of the SUS is based on the principles mentioned above, but also on its organizational model. Thus, it is divided into levels of care capable of strategically meeting the complexities of health, in order to be organized into Primary Health Care (PHC), Secondary Health Care and Tertiary Care. However, PHC represents the main sphere to guarantee the right to health, especially because it is, among the others, the context responsible for about 80% of health demands and, consequently, of the decision-making processes involving health care for the population, in order to reduce the flow of illnesses and injuries of patients destined for Tertiary Care, especially with regard to surgical processes and hospitalization (3).

Thus, in the context of the National Primary Care Policy - PNAB (Brasil, 2017), PHC encompasses actions aimed at prevention, protection, diagnosis, treatment, rehabilitation, harm reduction, palliative care, health promotion and surveillance, developed by multiprofessional teams (4). Thus, through the work developed by family doctors, nurses specialized in community health, community agents, and other trained professionals, this approach integrates interdisciplinary competencies and intersectoral strengths to address the social determinants of health. In addition to being resolute, PHC intercepts demands and offers effective responses, consolidating itself as the ideal model for the SUS (5).

Therefore, similar to other health systems, the functioning of the SUS depends fundamentally on the ability to promote health, as well as on the care of sick people who require professional care; thus to be effective, the system is subordinated to a qualified professional body, in line with the understanding that its primary axis is human capital. Despite this reality, nursing is the largest workforce in the health area and, consequently, in SUS and health systems worldwide(3).

International Nursing Institutions, such as the International Council of Nurses (ICN) and national bodies linked to it, such as the Federal Nursing Council (COFEN) in Brazil, as well as global health organizations, such as the World Health Organization (WHO), advocate the strengthening of health systems, granting professional autonomy to nurses to qualify and extend the reach of care practices (6–11). Thus, the Region of the Americas, since 2014, through the Pan American Health Organization (PAHO), has advocated Advanced Nursing Practice (APN) as a strategy for complying with its resolutions (10,12–13). This aspect represents the expansion of the scope of professional practice, with interventions based on the best evidence, patient profile, professional skills, technological resources, and appropriate legislation that, together, through robust protocols, may be able to produce efficient health outcomes for individuals, families, and communities served by accredited nurses (14–17).

Considering the APN as an effective strategy to address the challenges in access, continuity and efficiency in the provision of primary health care, the Organization for Economic Co-operation and Development (OECD) analyzed its development in five countries integrated to it, which are at different stages of its implementation: two countries (United States and Canada) that have a long experience in this process, Australia, which has moderately long experience, as well as France and Italy, which are still in the initial phase of implementing the APN in PHC. The evolution observed in the study shows a growing disparity between the countries that led this practice, in which there was an expansion of the APN, while those that adopted it only recently are still in the early stages, or still debating its adoption. In leading countries, this practice has been shown to be effective in expanding access to PHC, continuity of care, reducing hospital readmissions and greater patient satisfaction, without prejudice to the quality or safety of care when properly trained (18). It is important, however, to consider the specificities, potentials and strengths of the APN in the largest universal health system, notably in the context of PHC, based on the SUS.

From this perspective, numerous and important initiatives have been taken, both in terms of interventions in care practice, in academic spaces and in associative, regulatory and policy-making contexts (^8, 19–21^). In view of the above, it is worth asking: what is the panorama of the EPA in alignment with the context of PHC when it comes to the SUS?

Thus, motivated by the interest in contributing to the understanding of the evolutionary trajectory of APS in Brazilian PHC, we propose to investigate the ANP competencies that PHC nurses already develop in the country and to present evidence that contributes to the intensification of the debate on this theme and constitutes an incentive and subsidy for the implementation of ANP in Brazil.

### Objective

To categorize Advanced Nursing Practice activities in Brazil, registered in the Health Production reporting platform of the Health Information System for Primary Care, by Primary Health Care nurses.

## METHOD

### Ethical Aspects

This study is exempt from the appreciation of the Research Ethics Committee (REC) linked to the CEP/Conep System, since it uses exclusively secondary data, without nominal identification of the participants. The data analyzed is in the public and unrestricted domain, with no implications related to privacy, security or access control.

### Study Design, Period, and Location

This is a documentary study (^22^), exploratory and nationwide, conducted in 2024, based on public data available on websites linked to the Ministry of Health in Brazil, reference year in 2023 (^23–25^).

### Data Source

Data were extracted from the SISAB Health/Production Reporting Platform (^23^). In the Brazilian context, the National Primary Care Policy (PNAB) was reestablished in 2017, providing guidelines for the organization of Primary Care (PHC), one of the components of the Health Care Network. The document terminologically equates PHC to PHC, defined as a set of individual, family, and collective health actions, covering promotion, prevention, protection, diagnosis, treatment, rehabilitation, harm reduction, palliative care, and health surveillance, as provided for in Article 2 of the PNAB (^4^).

The PNAB defines, in articles 6 and 7, as well as in its Chapter I and annexes, the responsibilities common to health professionals in PHC. Specifically, item 4.2.1 details the nurse’s duties. In addition, SISAB is a public system used for information management in the SUS, allowing the collection of production data from PHC professionals. The system captures information entered via Simplified Data Collection, Electronic Citizen Record and mobile applications, such as e-SUS Territory and Collective Activity.

The SISAB data reflect the actions carried out by nurses and other professionals in PHC. The municipalities send monthly reports to the system, containing data on production and collective activities. This process ensures that nursing practices are aligned with the PNAB guidelines.

### Data Collection

Data were collected between June 23 and 27, 2024, following a protocol previously prepared by the authors. This protocol comprised systematic steps to ensure the integrity and reproducibility of the collection.

The ICN defines twenty-one characteristics of the Advanced Practice Nurse (ANP), distributed in three domains: Educational Preparation, Nature of Practice and Advanced Practice Nursing. This global nursing guiding document was used to relate the characteristics of advanced practice nursing converging with the nurses’ competencies established by the PNAB, according to the guidelines for the organization of Primary Care, within the scope of the SUS.

Thus, for each characteristic recommended by the ICN, the correspondence with the guidelines established in the PNAB, in its Chapter I and annexes, was identified. Thus, a comprehensive analysis of this policy was carried out, considering the main responsibilities attributed to nurses within the scope of PHC.

SISAB brings together the specific actions carried out by nurses and other professionals in the scope of primary care. The records are processed by the municipalities, which send monthly reports to SISAB, such as production data and collective activities. This process ensures that nurses’ practices are aligned with the guidelines of the National Primary Care Policy (PNAB).

Thus, through the extraction of data from SISAB, information on the production of nurses and reports of indicators by state, municipality, health region and team were obtained. To identify and categorize the specific actions performed by nurses within the scope of PHC, the codes present in the SISAB reporting platform were mapped in the Health/Production and Health/Collective Activity categories.

Procedures and care existing in the Management System of the Table of Procedures, Medications and Orthoses, Prostheses and Special Materials of the SUS (SIGTAP) that correspond directly to the expanded clinical activities performed by nurses were considered. The sequence of searches, by filter, carried out according to the illustration in Figure 1, made it possible to obtain a set of data, which were analyzed and related to the minimum competencies that characterize the APN, described in the “*Advanced Practice Nursing Characteristics Guidelines on Advanced Practice Nursing 2020 of the International Council of Nurses”,* distributed in three domains and 21 items.

**Figure 1.**
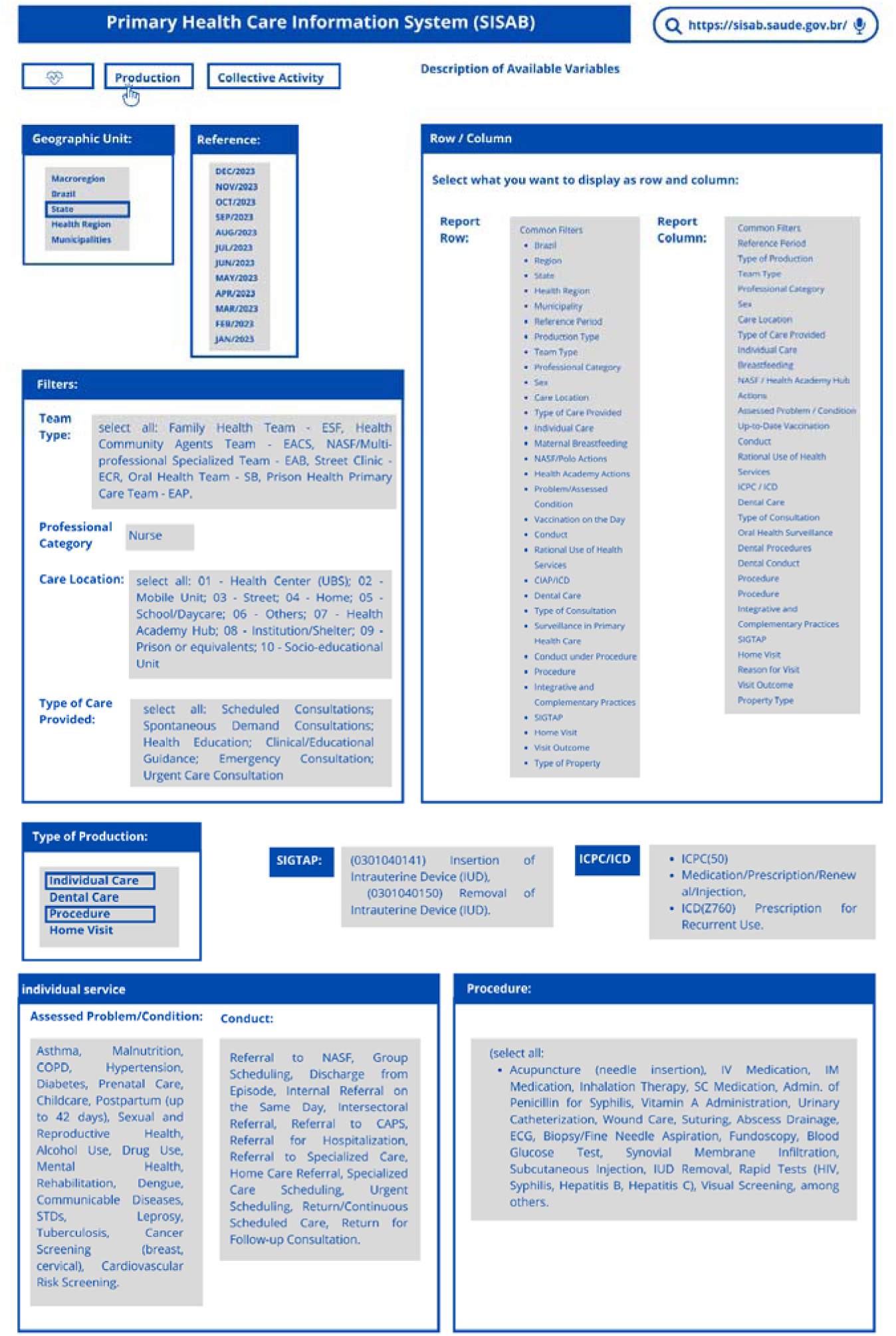
Illustrated sequence of search codes, by filter, to identify the activities performed by PHC nurses. Ribeirão Preto, São Paulo, Brazil. Source: SISAB (2024); illustration made by the authors.

In the results session, the data were organized in three tables, in which the minimum competencies of the ICN guided the indication of correspondence with the responsibilities of the nurse, contained in the 2017 PNAB guidelines of the Ministry of Health of Brazil, and APN activities registered in SISAB. These codes were organized in a table to compose the third column, allowing the identification and categorization of the specific actions performed by these professionals within the scope of PHC.

In the stage of mapping the codes present in the SISAB reporting platform, specific filters were used to extract data in the Health/Production and Health/Collective Activity categories. Fig 1

### Data Analysis

To identify and categorize the specific actions performed by nurses within the scope of PHC, the codes present in the SISAB reporting platform were mapped, in the Health/Production and Health/Collective Activity categories.

Procedures and care existing in the unified table of SIGTAP that correspond directly to the expanded clinical activities performed by nurses were considered. The sequence of searches, by filter (carried out according to the illustration in Figure 1), made it possible to obtain the raw data.

Comparative analysis was used on the data, relating them to the minimum competencies that characterize Advanced Nursing Practice, described in the “*Advanced Practice Nursing Characteristics Guidelines on Advanced Practice Nursing 2020 of the International Council of Nurses” of* the ICN), distributed in three domains, namely: Educational Preparation, Nature of Practice and Advanced Practice Nursing and 21 items/characteristics, described in their respective domains (Tables 1, 2 and 3).

Thus, the analysis was based on the aforementioned ICN document, as a global guide for nursing, to relate the characteristics of Advanced Practice Nursing converging with the competencies of nurses established by the PNAB, according to the guidelines for the organization of Primary Care, within the scope of the SUS. Thus, a comprehensive analysis of this policy was carried out, considering the main responsibilities attributed to nurses within the scope of PHC in accordance with the codes mapped on the SISAB reporting platform.

## RESULTS

The comparative analysis between the ICN and PNAB guidelines resulted in the classification presented in the third column of Chart 1, which reveals the unavailability of corresponding interventions performed by nurses in PHC, registered in SISAB.

**Chart 1.** Matrix for the analysis of competencies and interventions of Advanced Practices of Nurses in Primary Health Care, classified according to the Educational *Preparation domain*, according to the guidelines of the International Council of Nurses, National Primary Care Policy and the Ministry of Health of Brazil. Ribeirão Preto, São Paulo, Brazil.

| Educational Preparation |  |  |
| --- | --- | --- |
| International Council of Nurses, 2020 | National Primary Care Policy 2017 | Health Information System for Primary Care 2023 |
| 1. Additional educational preparation to training as a generalist or specialist nurse, with a minimum requirement of a full master's programme (master's level modules taken as separate courses do not meet this requirement). | Nurse, preferably a specialist in family health (currently, diverges from the International Council of Nurses, which recommends at least a master's degree in Advanced Nursing Practice. | unavailable |
| 2. Formal recognition of educational programs that prepare nurses specifically for Advanced Practice Nursing (e.g., accreditation, approval, or authorization by governmental or nongovernmental bodies). | CHAPTER I<br>RESPONSIBILITIES<br>VII - articulate with the Ministry of Education strategies to induce curricular changes in undergraduate and graduate courses in the health area, aiming at the training of professionals and managers with an adequate profile for Primary Care | unavailable |
| 3. A formal system of accreditation linked to defined educational qualifications. | In Brazil, the accreditation of nurses in Primary Health Care is obtained through the National Registry of Health Establishments, after professional registration with the category's class council. | unavailable |

Comparing the correspondence between the guidelines of the ICN and PNAB (Chart 1), it is verified that the three items that make up the dimension of *professional preparation* are present in the PNAB, but with a lower requirement than the ICN, as it requires specialization in Family Health instead of a Professional Master’s Degree in APN; in addition, although the PNAB is responsible for articulation between the Ministry of Health and the Ministry of Education for the adoption of strategies that induce change curricula for professional training, with management competencies in favor of PHC, it should be noted that in Brazil, the accreditation of PHC nurses is obtained through the CNES (National Registry of Health Establishments), upon proof of professional registration with the class council. Differently, the ICN recognizes ANP accreditation for the portability of a master’s degree in APN.

It should also be noted that the three items that make up professional training are not covered by the Ministry of Health in its PHC system.

In relation to the ten components of the “Nature of Practice” dimension (Chart 2), the essential differentiation is in the level of complexity, compatible with a APN when, in the case of this study, we are verifying expanded practices at the PHC level, which is especially observed in the interventions contained in items five, nine and ten of Chart 2, about the APN activities performed by the Nurse in PHC, according to SISAB code records, in Brazil in 2023.

**Chart 2.** Matrix of analysis of competencies and interventions of Advanced Practices of Nurses in Primary Health Care, classified according to the *Nature of Practice domain,* according to the guidelines of the International Council of Nurses, National Primary Care Policy and the Ministry of Health of Brazil. Ribeirão Preto, São Paulo, Brazil.

| Nature of the Practice |  |  |
| --- | --- | --- |
| International Council of Nurses, 2020 | National Primary Care Policy 2017 | Health Information System for Primary Care 2023 |
| 4. Role assigned to actions focused on the care (direct or indirect), prevention, and cure of disease in advanced-level health services, including rehabilitation care and chronic disease management. This role is beyond the scope of a generalist or specialty nurse's practice. | 4.2. The specific attributions of the professionals of the teams that work in Primary Care are:<br>4.2.1 - Nurse:<br>I - Provide health care to individuals and families linked to the teams and, when indicated or necessary, in all life cycles;<br>II - Perform nursing consultations; | Consultations<br>Problem/Condition Assessed: Asthma, Malnutrition, Diabetes, Chronic Obstructive Pulmonary Disease, Hypertension, Obesity, Prenatal Care, Childcare, Puerperium (up to 42 days), Sexual and Reproductive Health, Smoking, Alcohol User, Other Drug User, Mental Health, Rehabilitation, Communicable Diseases: Dengue, Sexually Transmitted Disease, Leprosy, Tuberculosis; Breast and cervical cancer screening; Cardiovascular risk screening. |
| 5. Ability to manage full episodes of care and complex health issues, including vulnerable, limited-access, and at-risk populations. | II - Perform nursing consultations, procedures, request complementary exams, prescribe medications according to protocols, clinical and therapeutic guidelines, or other technical regulations established by the federal, state, municipal or Federal District manager, observing the legal provisions of the profession;<br>III - Perform and/or supervise reception with qualified listening and risk classification, according to established protocols; | Interventions: Chemical cauterization of small lesions, Nail surgery (canthoplasty); Stoma care, Special dressing; Abscess drainage; Cardiogram; Diabetic foot examination; Excision/biopsy/puncture of tumor; Fundoscopy; Infiltration into synovial cavity; Subcutaneous foreign body removal; Removal of cerumen; Removal of a foreign body from the auditory and nasal cavity; Simple suture; Insertion of Intrauterine Device, Removal of Intrauterine Device. |
| 6. Ability to integrate research (evidence-based practice), education, leadership, and clinical management. | V - To carry out group activities;<br>VI - To plan, manage and evaluate the actions developed by nursing technicians/assistants, Community Health Agents and Endemic Disease Control Agents together with the other team members;<br>VII - Supervise the actions of the nursing technician/assistant and community health agent<br>VIII - Implement and keep updated routines, protocols and flows related to its area of competence in the (Basic Health Unit);<br>IX - To perform other duties in accordance with professional legislation, and which are of responsibility in their area of activity.<br>CHAPTER I<br>RESPONSIBILITIES<br>XVI - to guarantee adequate physical spaces and environments for the training of students and health workers, for in-service training and for permanent and continuing education in the Basic Health Units;<br>Art. 6 All health establishments that provide Primary Care actions and services, within the scope of the SUS, according to this ordinance, will be called Basic Health Units, all of which are considered potential spaces for education, training of human resources, research, in-service teaching, innovation and technological evaluation for the Health Care Networks, contained in its sole paragraph.<br>XIV - to promote the exchange of experiences between managers and workers, through horizontal cooperation, and to stimulate the development of studies and research that seek the improvement and dissemination of technologies and knowledge aimed at Primary Care; | Management and leadership:<br>Team meeting, Meeting with other health teams, Intersectoral meeting; Local Health Council, Control; Health education, Group care; Evaluation / collective procedure, Social mobilization and Permanent education actions; Topics for Meeting: - Administrative issues Work processes, Diagnosis of the territory; territory monitoring, Planning; monitoring of the team's actions, Case discussion;/ singular therapeutic project, Permanent education, Others. |
| 7. Broad and expanded autonomy (varies according to the country context and clinical setting). | III - Perform and/or supervise reception with qualified listening and risk classification, according to established protocols; (differs from International Council of Nurses, is restricted because it is dependent on protocols) | Autonomy according to the National Primary Care Policy: Diabetes; Hypertension; Prenatal; Sexual and reproductive health; Mental health; Communicable Diseases; Cancer screening; Tuberculosis; Leprosy; Gestational syphilis |
| 8. Case set management at an advanced level. | (not addressed in the National Primary Care Policy) |  |
| 9. Advanced assessment, judgment, decision-making, and diagnostic reasoning skills. | II - Perform nursing consultations, procedures, request complementary exams, prescribe medications according to protocols, clinical and therapeutic guidelines, or other technical regulations established by the federal, state, municipal or Federal District manager, observing the legal provisions of the profession;<br>III - Perform and/or supervise reception with qualified listening and risk classification, according to established protocols;<br>IX - To perform other duties in accordance with professional legislation, and which are of responsibility in their area of activity. | Spontaneous demand consultations and emergency care: Asthma, Malnutrition, Diabetes, Chronic Obstructive Pulmonary Disease, Hypertension, Obesity, Prenatal, Childcare, Puerperium (up to 42 days), Sexual and reproductive health, Smoking, Alcohol user, User of other drugs, Mental health, Rehabilitation, Communicable diseases: Dengue, Sexually Transmitted Disease, Leprosy, Tuberculosis; Breast and cervical cancer screening; Cardiovascular risk screening. |
| 10. Recognized advanced clinical skills that surpass those mastered by a generalist or specialized nurse. | IX - ditto above | Nurse consultation with autonomy for diagnostic evaluation and treatment, in addition to the execution of complex procedures, such as: Fundoscopy Chemical cauterization of small lesions; Nail surgery (canthoplasty); Fundoscopy; Excision/biopsy/puncture of tumor; Infiltration in cav. synovial; Simple suture; Insertion of intrauterine device, removal of intrauterine device. |
| 11. Ability to provide support and/or consulting services to other healthcare professionals, emphasizing professional collaboration. | V- Carry out group activities and refer users to other services, according to the flow established by the local network;<br>VI - To plan, manage and evaluate the actions developed by nursing technicians/assistants, community health agents, endemic disease control agents together with the other members of the team; | Management and leadership: Meeting with other health teams, Intersectoral meeting / local health council / control, Social mobilization. |
| 12. Plans, coordinates, implements and evaluates actions to improve health services at an advanced level. | I'm going to take action | Management and leadership:<br>Topics for Meeting: Diagnosis of the territory / monitoring of the territory, Planning / monitoring of the team's actions |
| 13. It is recognized as the first point of contact for clients and families (commonly, but not exclusively, in primary health care settings). | I - Provide health care to individuals and families linked to the teams and, when indicated or necessary, at home and/or in other community spaces (schools, associations, among others), in all life cycles; | It acts in a shared way in the user portfolio with the doctor, interspersing appointments in the agenda.<br>Health at School Program: Education, Health.<br>home/institutional visits by a higher education professional;<br>Home visit by a higher education professional. |

When comparing the types of advanced practice interventions provided for in the aforementioned Brazilian documents in relation to that of the ICN (Chart 3), with the exception of the last four (18 to 21), which are non-existent, the Brazilian documentation studied here gives nurses autonomy to diagnose and prescribe medications, diagnostic tests and therapeutic treatments, as well as to refer patients to other services and/or other professionals within the network.

**Chart 3.**
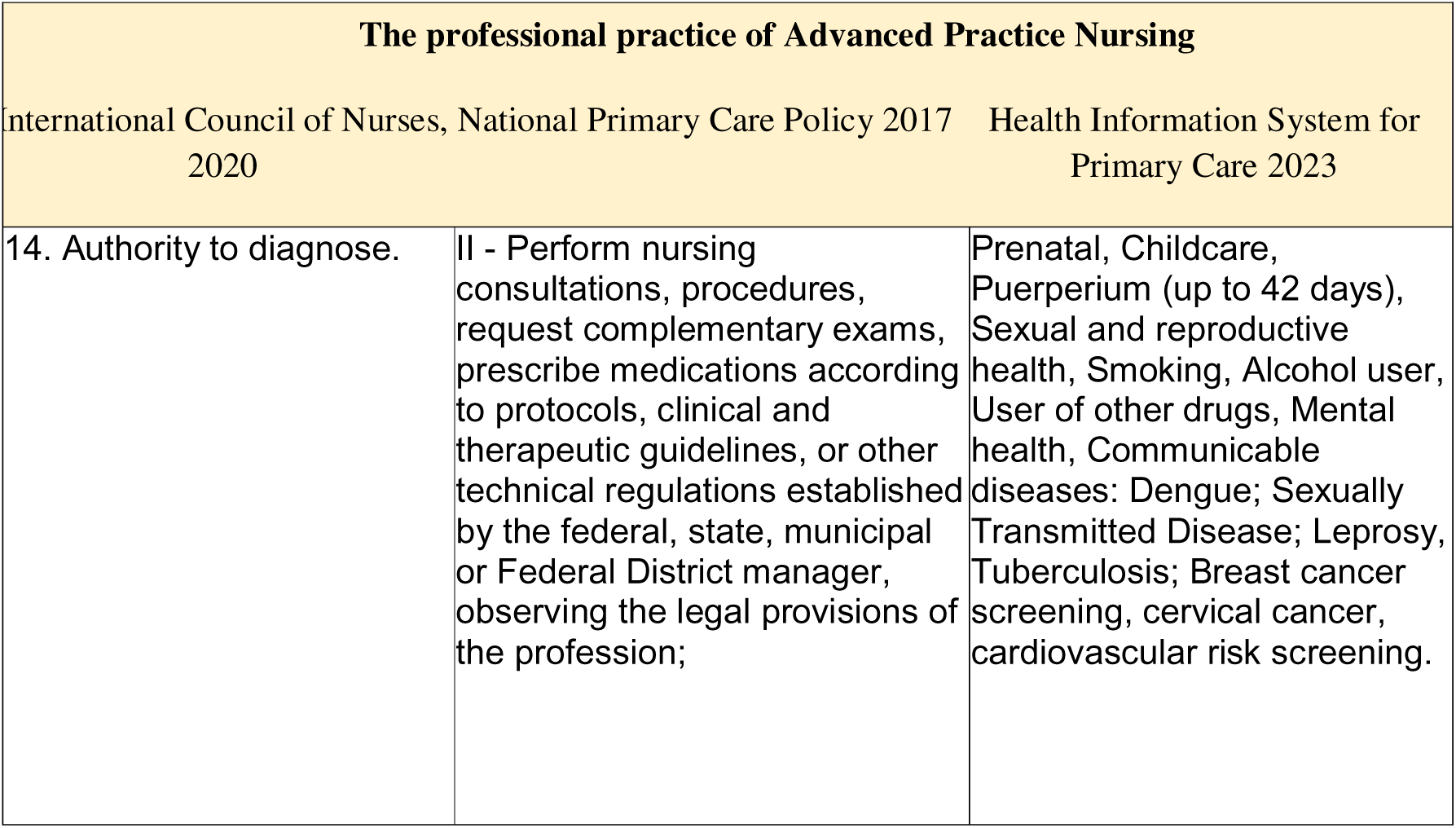
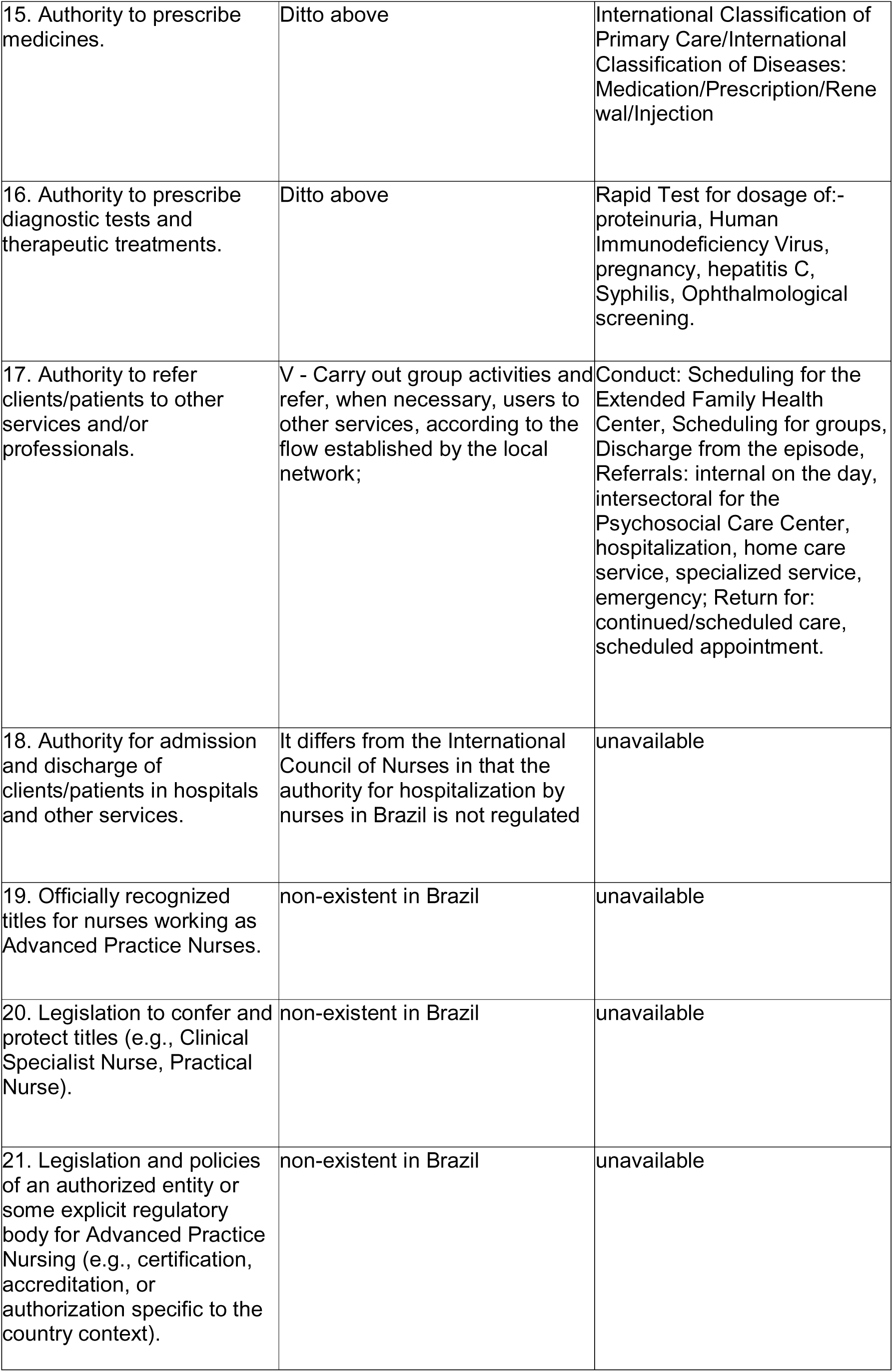
Matrix of analysis of competencies and interventions of Advanced Nursing Practices in Primary Health Care, classified according to the Advanced *Practice Nursing Exercise domain*, according to the guidelines of the International Council of Nurses, National Primary Care Policy and the Ministry of Health of Brazil. Ribeirão Preto, São Paulo, Brazil.

## DISCUSSION

Debates and continuous efforts by governments, under repeated recommendations from global health and economic organizations, have defended the power of primary care to rescue and transform health systems and ensure the quality of life of their peoples. PHC has been at the top of the global health policy agenda, deserving investments for its expansion since 1978, in Alma-Ata, to Astana, in 2018 (^26,27^).

In 2008, the World Health Report, marking the 60th Anniversary of the WHO and the 30th Anniversary of the Alma-Ata Declaration on Primary Health Care, recognizes unacceptable realities and avoidable setbacks in the performance of global health systems, which evidenced a movement against the comprehensive response to health needs, with inequity of access, high costs and erosion of trust in health systems, threatening social stability. Thus, the aforementioned report organized the reforms of Primary Health Care, in order to reorient health systems in favor of *health for all*; four sets of reforms were adopted:1) universal coverage reforms - for more equity in health, 2) service delivery reforms - to orient health systems to people, 3) leadership reforms—for more trusted health authorities, and 4) public policy reforms—to promote and protect community health (^28^).

From this perspective, the significant challenge of achieving “Health for all by the year 2000” is no longer a consensus, and has come to be seen by financial institutions as unattainable (^29,27^). Over 45 years, debates have been frequent in an attempt to implement health policies and strategies. However, goals were redefined and challenges were successively identified, including the shortage of human resources.

The shortage of health professionals is a global problem that directly impacts the quality of services and the ability of health systems to meet the demands of the population. This phenomenon is influenced by socioeconomic, political, and demographic factors that vary according to the context of each country or region. It is estimated that by 2030, the world will face a *deficit* of about 10 million health workers, mainly in low- and middle-income countries. This *deficit* affects, above all, nurses, doctors and community health agents. The shortage of health professionals generates negative impacts in several areas, such as: limitation in access to services, waiting time for care, limited coverage of services, and work overload of professionals. Strategies to reduce the *deficit* focus on expanding vocational training (^9^) and on the continuous training of human capital throughout working life, which is a real source for implementing change (^30^).

The 2020 State of Nursing report (^9^) brought more recent information on how to improve the nursing workforce around the world, highlighting professional value and pointing out what needs to be improved to improve health services and achieve global goals; above all, it stressed the need to invest considerably in: a) education of nurses, ensuring quality training; b) generation of jobs, offering better working conditions; c) development of leadership in the area, so that nurses can influence health policies.

International nursing organizations, consultants, and researchers have politically promoted the debate (^31–36^), some of them reiterating that the underutilization of nurses’ professional training (^34,37–39^) needs to be definitively overcome by a policy that invests in these professionals, ensuring that nurses can perform to their maximum potential and, thus, make a substantial contribution to improving global health and achieving sustainable development goals. Without the work of nurses, there will be no sustainable development and no universal health coverage for all, everywhere (^36^) The report also has the political strength capable of transforming the call into action, since it calls on Member States and other stakeholders to commit to this agenda of investments in nurses, highlighting that these human resources will not only boost health, but also contribute to advances in education, gender equality, decent work, and economic growth (^9^).

Thus, if there were already actions and evidence of positive results and recognition of the much-needed expansion of the scope of practice, giving nurses more autonomy, after the call for action in favor of investment in nursing by international bodies, involving ministries of health from 191 countries, it is clear that the APN is a vital and highly effective strategy to reduce scarcity and provide the achievement of universal health coverage.

ICN considers the power of nursing to take on advanced nursing practice (^40^), determining guidelines and offering concepts that can be adopted by policymakers that contemplate the use of the full potential of nurses, in the areas of government, education, health systems and professional practice to support advanced nursing practice initiatives. It is in this understanding that coherence and clarity in the conception of nursing capable of meeting the health needs of people and health systems globally are justified (^7^).

The APN is distinguished from traditional nursing practice by its level of specialization and professional autonomy, allowing nurses to perform complex interventions, prescribe medications (in some contexts), order diagnostic tests and develop therapeutic plans. In addition, these professionals play a key role in health education, team management, and public policy formulation.

Similarly, the convergence between the PNAB and ICN, seen in items 5, 6, 7, 9, 12 and 13 of this study, is in line with the findings in the recent scientific literature (^41–43^) when describing the professional autonomy of nurses in PHC with support in care protocols, which guarantee expanded clinics, decision-making, and increased problem-solving capacity centered on the user’s needs, continuity of care, construction of bonds with the community, autonomy in the areas of management/management of individual and collective practices aimed at prevention.

Similarly, in the Nature of Advanced Practice Nursing Practice and Practice domain, items 4, 7, 14, 15 and 16, PHC nurses in Brazil stand out for their clinical autonomy to diagnose, prescribe medications, diagnostic tests and therapeutic treatments to people with diabetes, hypertension, prenatal care, sexual and reproductive health, mental health, Communicable Diseases, Cancer Screening, Tuberculosis, Leprosy and Gestational Syphilis.

In 2021, APN actions in the context of the Family Health Strategy were mapped, highlighting the authority of nurses in requesting tests, such as clinical cytopathological analyses, screening mammography, blood count, fasting glucose, VDRL, HIV, HBsAg, urine culture, antibiogram, serology for toxoplasmosis, laboratory and imaging tests, in addition to fecal and urine tests, obstetric ultrasound, among other specific prenatal tests, women’s and children’s health. The same study also detected the autonomy to prescribe medications, such as folic acid, ferrous sulfate, nystatin, miconazole, fluconazole, paracetamol, antibiotics and other medications available in the nursing protocol of the city studied (^17^). In the same direction, in 2022, another study pointed out the frequency of nursing consultations, as well as the request for tests, such as blood count and other blood tests, ultrasound, as well as the prescription of medications, the most prescribed being ferrous sulfate and other supplements (44).

In an identical way, our study demonstrates in Items 5 and 10, the autonomous role of the nurse for diagnostic evaluation and treatment, in addition to the execution of complex procedures, such as; simple suture, fundoscopy, chemical cauterization of small lesions, insertion of Intrauterine Device (IUD) and others. It should be noted that, in 2023, the performance of simple sutures by nurses was regulated by COFEN (^45^).

After nurses were qualified for IUD insertion in 2018, it was found that until May 2021, 2,024 insertions were recorded by these professionals (^46^) in the state of Santa Catarina alone. At the national level, in that same year, nurses performed 4,653 IUD insertions, while physicians performed 13,590 (^47^). On the other hand, the number of 54,186 reproductive planning consultations was observed, of which 41,184 (76%) were by nurses and 13,002 (24%) by physicians. These findings highlight the importance of nurses in expanding the scope of action in PHC, evidencing their fundamental role in expanding access to services and health coverage.

Evidence such as these, in addition to others already recorded in the literature, associated with the analysis matrix produced in this study demonstrates that APN in the context of PHC is an undeniable reality in Brazil. Thus, it is to be expected that leading leaders from the regulatory, educational and health policy sectors, professional associations, scientific societies and nursing specialists, schools and colleges will transform the call into action, in their respective sectors, so that the evidence recorded here is reverted into actions for change.

As well emphasized, countries that wish to increase the scope of practice of nurses and the strengthening of the APN to improve access and coverage in health must ensure adequate regulation, standardization, remuneration, financing and training policies for these nurses capable of strengthening the quality of the health system, reducing costs, valuing nurses, favoring the problem-solving capacity of health care demands and boosting equity.

Nevertheless, Brazil still lacks deontological support that guarantees, in addition to the current legislation, possibilities for nurses to exercise the APN autonomously, since there are still normative and cultural barriers that distance nurses from their competences to the health demands of the population, as in the case of requesting exams, prescription of medicines filed and referrals, among others, in PHC.

In addition, the analysis matrix indicated here represents a comparative conceptual structure that demonstrates the evidence of advanced practices that have been assumed and recorded by nurses in the SUS data system, with open access.

### Study limitations

The documentary and exploratory nature of the study reveals potential limitations, especially when restricting the analysis of secondary data, without direct observation of the advanced practices performed by nurses. In addition, the exclusive use of SISAB may present information gaps, since the data depend on the quality of the registration carried out by each Brazilian municipality, whose coverage of national data does not guarantee homogeneity in the records among the different regions of the country, although this is the best possible indicator for a documentary study.

### Contributions to the field of Nursing

The matrix developed in this study represents a strategic management tool and the usability of the illustrated sequence of search codes, by filter, to identify the activities performed by PHC nurses, allows those interested to obtain data on nurses’ activities within the scope of PHC and, thus, to know the breadth of the scope of professional practice.

## CONCLUSION

The analysis matrix allowed us to categorize the activities of Advanced Nursing Practices in Brazil, in order to highlight the progress and limitations in the context of PHC, in a country of continental dimensions, which celebrates and reiterates universal access to health from the SUS.

In the study, it was observed that, although nurses have competencies that confer autonomy for important practices such as prescribing medications, requesting exams and referring patients, Brazilian policies have gaps in relation to the international recommendations of the ICN and the WHO itself.

The requirement of specialization in Family Health, instead of a master’s degree in APN, for example, evidences a certain mismatch in professional training, so as to compromise the standardization of the qualification required for nurses, at this level of training and intervention, for PHC. In addition, Brazil’s regulatory framework fully contemplates the components of the “Nature of Practice” of APN, limiting the expansion of these activities in PHC itself. However, this reality should be the object of debates and reflections on the demands and needs of Brazilian health and nursing.

## Data Availability

All data produced in the present study are available upon reasonable request to the authors

## Notes

### Competing Interest Statement

The authors have declared no competing interest.

### Funding Statement

This study was supported by the Brazilian Coordination for the Improvement of Higher Education Personnel (CAPES) Funding Code 001 the National Council for Scientific and Technological Development (CNPq) and the Federal Nursing Council (COFEn).

### Summary of Updates

I am submitting a revised version of the manuscript to correct the use of acronyms, adapting them from English (APN, EPA, PAE, ANP) to their appropriate equivalents in Portuguese, as required by the journal's guidelines. Thank you for your consideration.

